# Reduced Serum Hydrogen Sulfide Levels in Drug-Naïve Patients with Major Depressive Disorder: A Cross-Sectional Analytical Study from Eastern India

**DOI:** 10.64898/2026.05.03.26352330

**Authors:** Atrim Das, Pritam Datta, Nirmal Bera

**Author notes:** **Corresponding author:** Dr Nirmal Bera, Head of Department, Department of Psychiatry, North Bengal Medical College and Hospital, Darjeeling, West Bengal, India. **Contact —** Atrim Das / | Dr. Pritam Datta. **Conflict of interest:** The authors declare no conflict of interest. **Ethical approval:** The study was approved by the Institutional Ethics Committee of North Bengal Medical College and Hospital (Ref. No.: [ECR/1701/Inst/WB/22]). All procedures were conducted in accordance with the Declaration of Helsinki. Written informed consent was obtained from all participants prior to enrolment. **Data availability:** The data that support the findings of this study are available from the corresponding author upon reasonable request.

## Abstract

**Background:** Hydrogen sulfide (H₂S) is an endogenous gasotransmitter synthesised in the central nervous system (CNS) primarily by cystathionine β-synthase (CBS) and cystathionine γ-lyase (CSE). Pre-clinical studies consistently implicates H₂S deficiency in the pathophysiology of depression through disruption of synaptic plasticity, neuroinflammation, oxidative stress, and brain-derived neurotrophic factor (BDNF) signalling. Yet, we still lack direct clinical evidence quantifying circulating H₂S in patients with Major Depressive Disorder (MDD), particularly from South Asian populations. In this study, we measured serum H₂S levels in drug-naïve patients with MDD and compared them with healthy controls at a tertiary care center in eastern India. We examined the associations between serum H₂S and depression severity as assessed by the 17-item Hamilton Depression Rating Scale (HAM-D-17). This institution-based, cross-sectional analytical study was conducted at North Bengal Medical College and Hospital (NBMCH), West Bengal, India, over 12 months. Fifty drug-naïve patients fulfilling DSM-5 criteria for MDD and fifty age- and sex-matched healthy controls were enrolled by consecutive sampling. We quantified serum H₂S using the spectrophotometric methylene blue method and depression severity was assessed using HAM-D-17. Statistical analyses included independent-samples t-test, chi-square test, and linear regression. Serum H₂S was markedly and significantly lower in MDD patients (0.068 ± 0.044 µmol/L) compared with healthy controls (0.524 ± 0.272 µmol/L; p < 0.001), representing an approximately 7.7-fold reduction. HAM-D-17 scores were significantly higher in MDD patients (28.94 ± 12.78) than in controls (3.96 ± 2.31; p < 0.001). Linear regression across the combined cohort revealed a significant negative association between serum H₂S and HAM-D score (R² = 0.287; y = 24.64 − 26.84x; p < 0.001), indicating that higher serum H₂S was associated with lower depression severity. Within the MDD group alone, the regression was weak (R² = 0.061), consistent with a floor effect. Within the control group alone, the regression was strong (R² = 0.772). No significant associations were found between serum H₂S and any sociodemographic variable in either group. Drug-naïve MDD patients exhibited substantially reduced serum H₂S levels compared with healthy controls, and lower H₂S was associated with greater depression severity. These findings provide direct clinical evidence from an Indian population supporting the H₂S deficiency hypothesis of depression and suggest that the CBS/CSE–H₂S axis may represent a novel biomarker and therapeutic target in MDD.

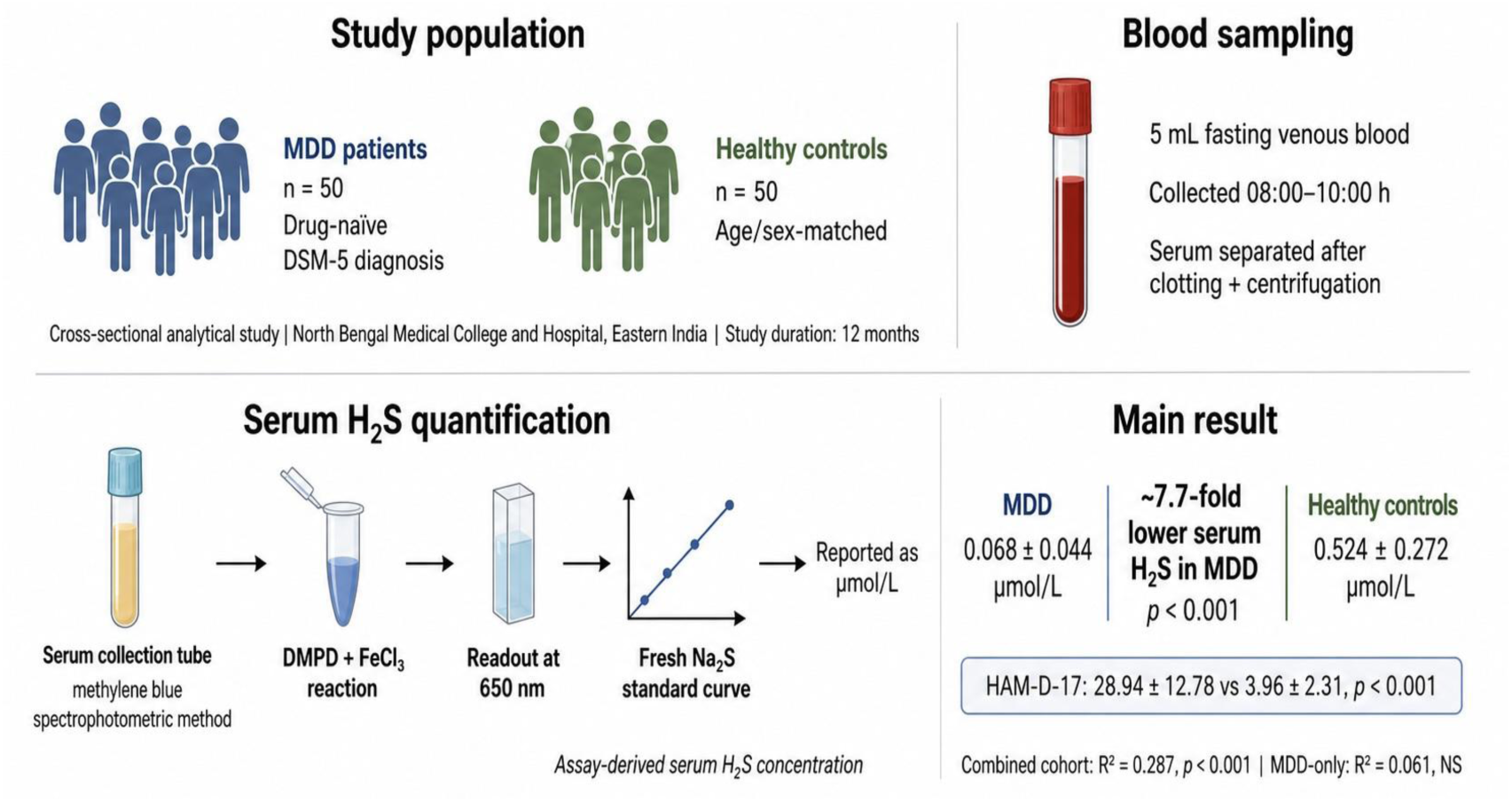

## 1. Introduction

Major Depressive Disorder (MDD) is one of the most prevalent and disabling psychiatric conditions worldwide, affecting an estimated 280 million people and accounting for the largest share of years lived with disability among all mental disorders [1]. Despite decades of research and the availability of multiple pharmacological treatments, around an estimated 30 to 50% of patients fail to achieve full remission with first-line antidepressants [2]. This persistent therapeutic gap underscores the need to identify novel neurobiological mechanisms that could serve as both biomarkers and therapeutic targets in MDD. The monoamine hypothesis of depression, which has dominated the field for over half a century, is increasingly recognised as incomplete. Emerging evidence implicates a broader set of pathophysiological processes in MDD, including neuroinflammation, oxidative and nitrosative stress, impaired synaptic plasticity, neurotrophic factor depletion, mitochondrial dysfunction, and dysregulation of the hypothalamic-pituitary-adrenal (HPA) axis [3]. Within this evolving framework, gasotransmitters, which are endogenously produced gaseous signaling molecules have attracted growing attention as pleiotropic regulators of CNS function whose dysregulation may contribute to depression. Hydrogen sulfide (H₂S) is now the third endogenous gasotransmitter to be characterised, alongside nitric oxide (NO) and carbon monoxide (CO) [4]. In the CNS, H₂S is produced enzymatically by three principal pathways (Figure 1): (i) cystathionine β-synthase (CBS), which is highly expressed in astrocytes and neurons of the cortex and hippocampus and represents the dominant source of brain H₂S, (ii) cystathionine γ-lyase (CSE), which is expressed in spinal cord microglia and cerebellar granule neurons, and (iii) 3-mercaptopyruvate sulfurtransferase (3-MST) acting in concert with cysteine aminotransferase (CAT), which generates H₂S within mitochondria [5, 6]. CBS activity is positively regulated by S-adenosylmethionine (SAM) and negatively regulated by carbon monoxide, whereas CSE is regulated by intracellular calcium and transcription factors. 3-MST is regulated by calcium and redox state [5].

**Figure 1:**
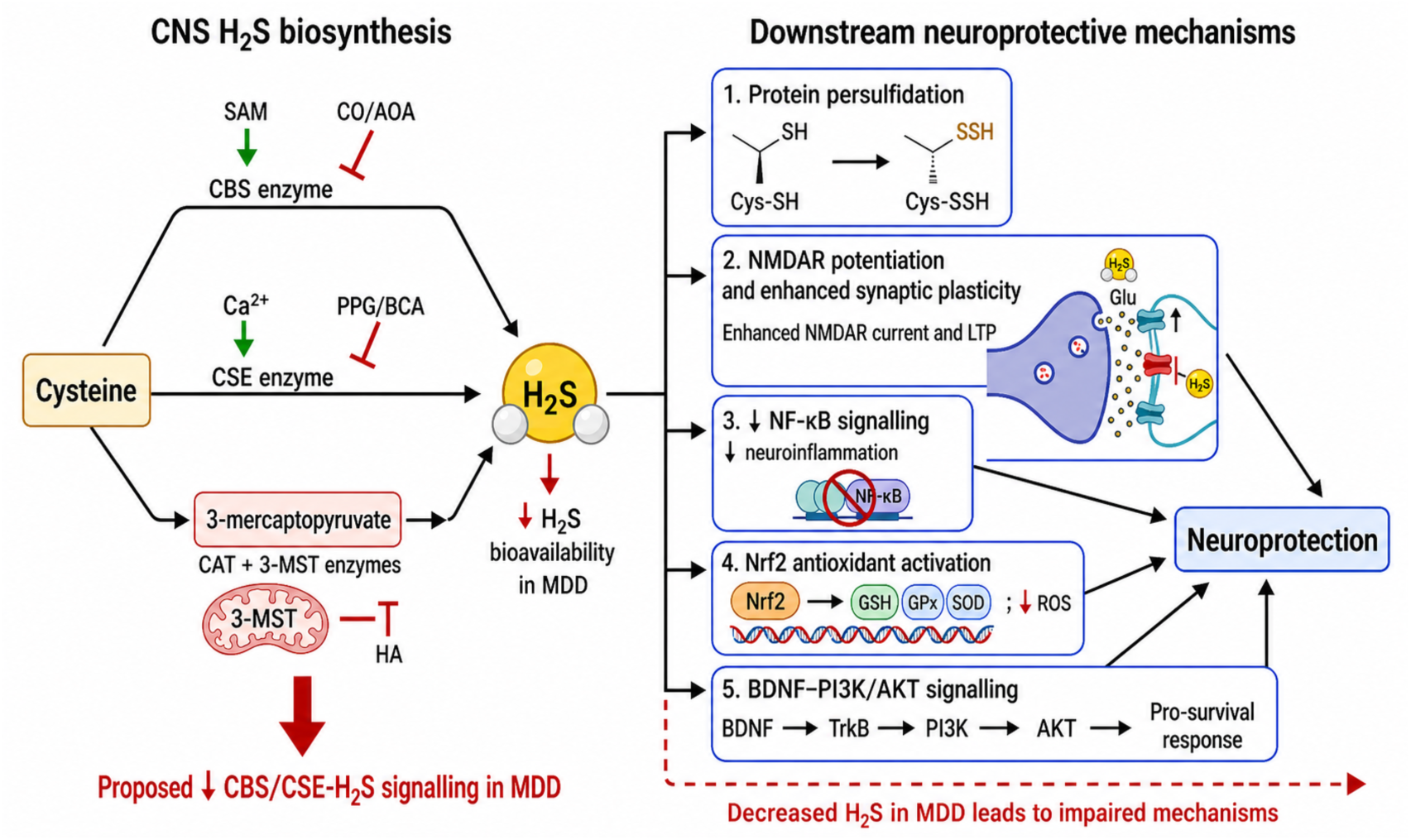
The major enzymatic pathways generating H₂S in the CNS and their downstream neuroprotective effects. Schematic illustrating the three enzymatic pathways of H₂S biosynthesis in the CNS: (i) CBS (cystathionine β-synthase), the dominant brain H₂S source, regulated by SAM (activator) and CO/ AOA (inhibitors); (ii) CSE (cystathionine γ-lyase), regulated by Ca²⁺ and inhibited by PPG/BCA; and (iii) 3-MST + CAT (mitochondrial pathway), inhibited by HA. All three pathways converge to produce H₂S, which exerts downstream neuroprotective effects through protein persulfidation, NMDAR potentiation, NF-κB suppression, Nrf2 activation, and BDNF/PI3K/ AKT upregulation. In MDD, CBS/CSE activity is reduced, leading to H₂S deficiency and downstream impairment of these neuroprotective mechanisms. CBS = cystathionine β-synthase; CSE = cystathionine γ-lyase; 3-MST = 3-mercaptopyruvate sulfurtransferase; CAT = cysteine aminotransferase; SAM = S-adenosylmethionine; AOA = aminooxyacetic acid; PPG = propargylglycine; BCA = β-cyanoalanine; HA = hydroxylamine.

**Figure 2:**
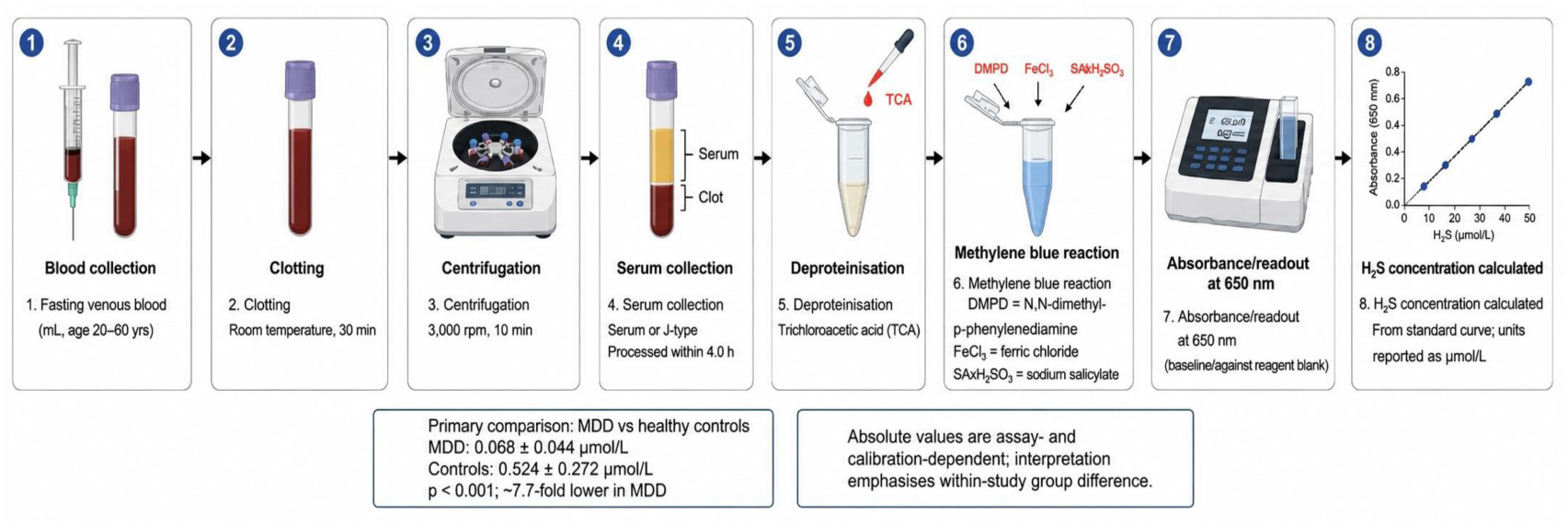
Serum H₂S assay workflow.

H₂S exerts its biological effects through several mechanisms that are directly relevant to the neurobiology of depression. The primary post-translational mechanism is protein persulfidation (S-sulfhydration), in which H₂S adds a sulphur atom to reactive cysteine residues, modifying protein activity and downstream signalling [5]. Through this mechanism, H₂S enhances N-methyl-D-aspartate receptor (NMDAR) currents and promotes hippocampal long-term potentiation (LTP), which is a cellular correlate of learning and memory via CaMKII activation and D-serine release from astrocytes [5, 6]. H₂S also activates the Nrf2 antioxidant pathway by persulfidating Keap1, upregulating cytoprotective genes including those involved in glutathione (GSH) synthesis [5]. Through persulfidation of the p65 subunit of NF-κB, H₂S suppresses pro-inflammatory transcriptional programmes and inhibits the NLRP3 inflammasome, reducing microglial activation and pro-inflammatory cytokine production [5, 6]. Additionally, H₂S activates the PI3K/AKT/mTOR pro-survival pathway, upregulates BDNF and its receptor TrkB, and modulates Sirt1 and Sirt6, that are deacetylases regulating neuronal survival, energy metabolism, and anti-inflammatory responses [6]. Collectively, these mechanisms position H₂S as a pleiotropic neuroprotective molecule whose deficiency could contribute to the oxidative stress, neuroinflammation, synaptic dysfunction, and neurotrophic factor depletion that characterise MDD. Preclinical evidence strongly supports the H₂S deficiency hypothesis of depression. In a chronic unpredictable mild stress (CUMS) mouse model, hippocampal H₂S levels and CBS expression were significantly reduced, accompanied by decreases in BDNF, synaptophysin (SYN), and postsynaptic density protein 95 (PSD-95), all of which are markers of synaptic integrity [7].

Treatment with SAM, a CBS agonist, restored hippocampal H₂S and CBS levels, reversed synaptic ultrastructural deficits, and ameliorated depression-like behaviours in sucrose preference, open-field, and forced swim tests [7]. In a lipopolysaccharide (LPS)-induced depression model, plasma H₂S and GABA levels were significantly reduced, hippocampal CA1 neuron counts decreased, NF-κB phosphorylation increased, and SYN and PSD-95 expression fell. Administration of a novel multifunctional H₂S and GABA donor (BGS) reversed all these changes [8]. A comprehensive systematic review of 14 preclinical studies published between 2021 and 2025 confirmed that exogenous H₂S administration, most consistently via NaHS exerts antidepressant and anxiolytic effects across diverse depression models (CUMS, LPS, corticosterone, PTSD, diabetic depression, neuropathic pain) through multiple converging pathways including PI3K/AKT/mTOR, Sirt1/Sirt6, NF-κB/NLRP3, cGAS-STING, CREB/BDNF, and ER stress suppression [6]. Despite this compelling preclinical evidence, direct clinical measurement of circulating H₂S in MDD patients remains limited. Yang et al. (2021) conducted the first dedicated clinical study, measuring plasma H₂S in 47 depressed patients and 51 healthy controls in China using a sensitive monobromobimane (MBB) fluorometric method, and found significantly lower H₂S in depressed patients with a severity-dependent gradient [9]. Dutta et al. (2024) subsequently reported significantly reduced serum H₂S in patients with major depressive episodes in the context of bipolar disorder (MDP) at a tertiary centre in West Bengal, India, using the methylene blue spectrophotometric method, and demonstrated excellent diagnostic accuracy (AUC = 0.964) [10]. A recent systematic mini-review by Pinna et al. (2025) synthesised this clinical evidence alongside the preclinical literature and identified the need for larger, independent clinical datasets from diverse populations [6]. To our knowledge, no prior study has quantified serum H₂S specifically in drug-naïve patients with unipolar MDD from the Indian subcontinent and correlated it with depression severity.

The drug-naïve design is critical since antidepressants, particularly SAM-based nutraceuticals, may directly modulate CBS activity and H₂S levels, confounding any observed association. The present study was therefore designed to: (i) measure serum H₂S levels in drug-naïve MDD patients and healthy controls; (ii) compare H₂S levels between the two groups; and (iii) examine the relationship between serum H₂S and HAM-D-17 depression severity scores.

## 2. Materials and Methods

### 2.1 Study Design and Setting

This was an institution-based, cross-sectional analytical study conducted at the Department of Psychiatry and the Department of Biochemistry, North Bengal Medical College and Hospital (NBMCH), Darjeeling, West Bengal, India. It is a tertiary care teaching hospital serving a predominantly rural population in the sub-Himalayan region of eastern India. The study was conducted over a period of 12 months.

### 2.2 Participants

#### Patient group

We enrolled fifty consecutive drug-naïve patients aged 18 to 60 years who fulfilled DSM-5 criteria for a current episode of Major Depressive Disorder [11] from the Psychiatry outpatient and inpatient services. “Drug-naïve” was defined as no prior or current exposure to any psychotropic medication, including antidepressants, antipsychotics, mood stabilisers, or anxiolytics. This criterion was verified by direct patient and caregiver interview and review of available medical records.

#### Control group

Fifty healthy volunteers, matched to the patient group for age (within 5 years) and sex, were recruited from hospital staff, attendants, and community members. Controls had no personal or first-degree family history of any psychiatric disorder and no current medical illness.

#### Inclusion criteria (patients)

Age 18–60 years; DSM-5 diagnosis of MDD confirmed by a consultant psychiatrist; drug-naïve status; willingness to provide written informed consent.

#### Inclusion criteria (controls)

Age 18–60 years; no current or lifetime psychiatric diagnosis; no psychotropic medication; no significant medical illness; willingness to provide written informed consent.

#### Exclusion criteria (both groups)

We excluded anyone with significant medical or surgical illness including hepatic, renal, thyroid, or cardiovascular disease, pregnancy or lactation, substance use disorder (other than nicotine), bipolar disorder, psychotic disorder, or other DSM-5 psychiatric comorbidity. Finally we ruled out anyone using any medication known to affect H₂S metabolism (e.g., N-acetylcysteine, SAM, homocysteine-lowering agents, NSAIDs).

### 2.3 Ethical Approval

The study was approved by the Institutional Ethics Committee (IEC) of North Bengal Medical College and Hospital, Darjeeling (IEC Memo No.: IEC/NBMC/M-07/069/2023, dated 26 May 2023; IEC Registration No.: ECR/1701/Inst/WB/2022). All procedures were conducted in accordance with the Declaration of Helsinki. Written informed consent was obtained from all participants prior to enrolment.

### 2.4 Sociodemographic and Clinical Assessment

A structured sociodemographic proforma was used to record age, sex, residence (rural/urban), language, religion, marital status, educational level, occupation, socioeconomic status (SES; Kuppuswamy scale [13]), family type, and duration of illness. Depression severity in the patient group was assessed using the 17-item Hamilton Depression Rating Scale (HAM-D-17) [12], administered by a trained psychiatrist. HAM-D-17 scores were categorised as: mild (8–13), moderate (14–18), severe (19–22), and very severe (≥23).

### 2.5 Blood Sample Collection and Serum H₂S Measurement

To keep natural daily fluctuations from skewing the data, fasting venous blood samples of 5 mL were collected from all participants under aseptic conditions between 08:00 and 10:00 hours to minimise diurnal variation. Samples were allowed to clot at room temperature for 30 minutes and centrifuged at 3,000 rpm for 10 minutes to obtain serum. Serum was stored at −20°C until analysis, with all samples processed within 48 hours of collection. Serum H₂S concentration was quantified using the spectrophotometric methylene blue method, as described by Siegel et al. and adapted for serum samples. Briefly, serum was deproteinised with trichloroacetic acid and reacted with N,N-dimethyl-p-phenylenediamine sulphate (DMPD) and ferric chloride (FeCl₃) in the presence of sulphide to form methylene blue. Absorbance was measured at 650 nm using a UV-visible spectrophotometer. A standard curve was constructed using sodium sulphide (Na₂S) solutions of known concentration, prepared fresh on the day of analysis. Results were expressed as µmol/L.

#### Methodological note on absolute H₂S values

The methylene blue spectrophotometric method measures primarily free and acid-labile sulphide fractions. Absolute H₂S values obtained by this method are sensitive to sample handling, deproteinisation conditions, standard curve preparation, and the specific Na₂S calibration stock used. As noted in the recent literature, there is substantial inter-laboratory variability in absolute H₂S values even when the same nominal method is used [6]. The values reported in this study (controls: 0.524 ± 0.272 µmol/L) reflect the specific calibration conditions of our laboratory and should be interpreted in the context of the within-study fold-difference between groups (∼7.7-fold) rather than compared directly with absolute values from other laboratories. The large and statistically robust between-group difference (p < 0.001) and the significant correlation with HAM-D severity are the primary findings of this study.

### 2.6 Statistical Analysis

Data were entered and analysed using IBM SPSS Statistics version 20. Continuous variables are presented as mean ± standard deviation (SD). Categorical variables are presented as frequencies and percentages. Between-group comparisons of continuous variables were performed using the independent-samples t-test after confirming approximate normality by inspection of histograms and Q-Q plots. Categorical variables were compared using the chi-square test. Linear regression analysis was performed to examine the relationship between serum H₂S (independent variable) and HAM-D-17 score (dependent variable), both for the combined cohort (n = 100) and separately within each group (n = 50 each). Associations between serum H₂S and sociodemographic variables were assessed using chi-square tests after dichotomising H₂S at the median. A p-value of < 0.05 was considered statistically significant for all tests.

## 3. Results

### 3.1 Sociodemographic Characteristics

The sociodemographic profiles of the MDD patient group and the healthy control group are summarised in Table 1 and illustrated in Figure 3. The two groups were comparable in age distribution, with the majority of participants in both groups falling in the 21 to 30 year age bracket (MDD: 46%; controls: 40%). The MDD group was predominantly female (74%; n = 37) compared with the control group (60%; n = 30). The majority of MDD patients were from rural areas (78%; n = 39) compared with 56% (n = 28) of controls. In terms of marital status, a notably higher proportion of MDD patients were single (62%; n = 31) compared with controls (46%; n = 23), while 44% (n = 22) of controls were married compared with only 20% (n = 10) of MDD patients. Illiteracy was more prevalent in the MDD group (50%; n = 25) than in controls (20%; n = 10). Unskilled labour was the most common occupation in the MDD group (30%; n = 15). The mean duration of illness in the MDD group was 19.26 ± 10.63 months and the mean duration of follow-up in the control group was 18.32 ± 10.32 months. No statistically significant differences were found between the two groups in age, religion, language, family type, or socioeconomic status (all p > 0.05).

**Figure 3:**
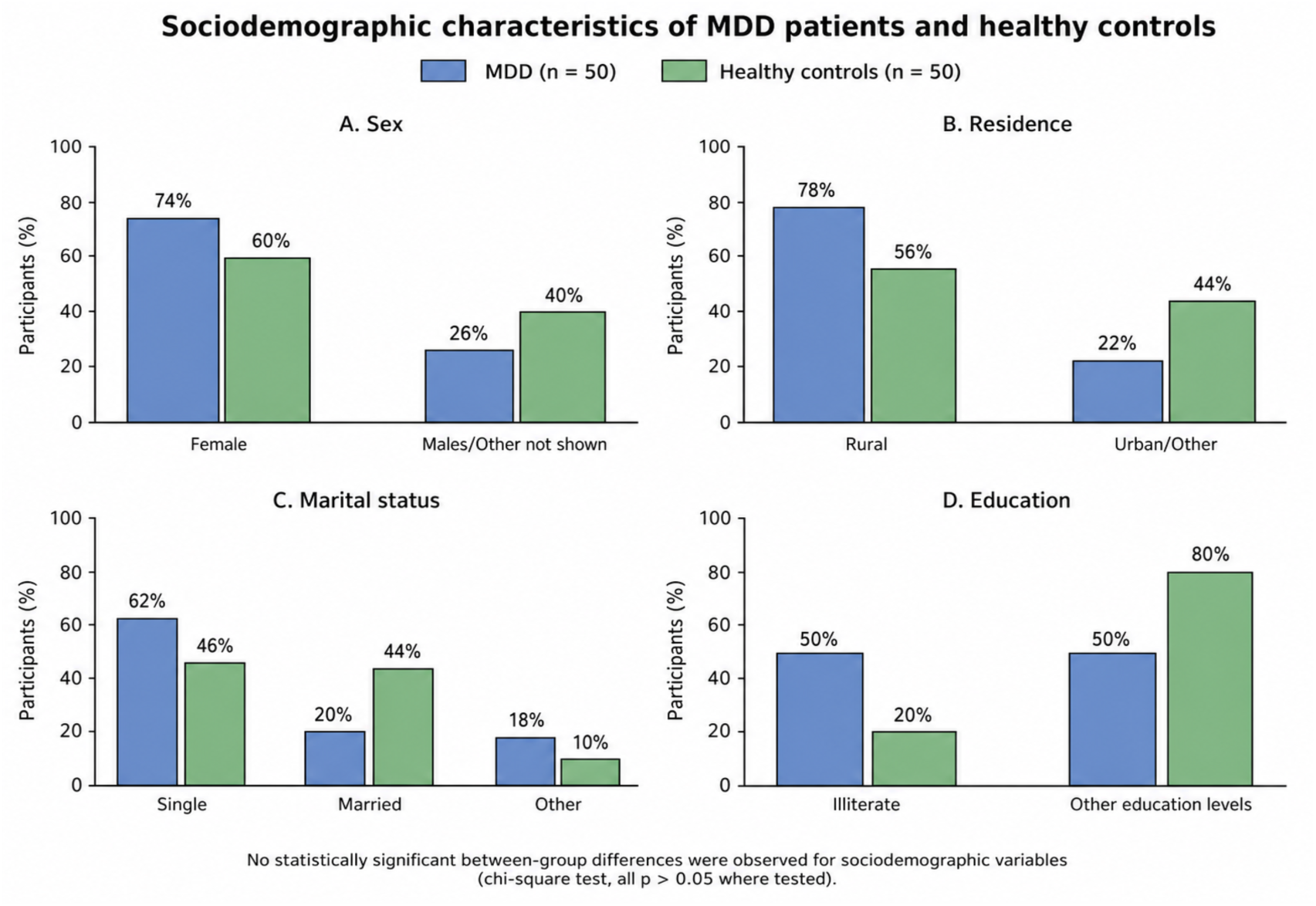
Sociodemographic characteristics of MDD patients and healthy controls.

**Table 1.** Sociodemographic characteristics of MDD patients and healthy controls.

| Variable | MDD (n = 50) | Controls (n = 50) | p-value |
| --- | --- | --- | --- |
| Age group: 21-30 years, n (%) | 23 (46%) | 20 (40%) | NS |
| Sex: Female, n (%) | 37 (74%) | 30 (60%) | NS |
| Residence: Rural, n (%) | 39 (78%) | 28 (56%) | — |
| Marital status: Single, n (%) | 31 (62%) | 23 (46%) | — |
| Marital status: Married, n (%) | 10 (20%) | 22 (44%) | — |
| Education: Illiterate, n (%) | 25 (50%) | 10 (20%) | — |
| Occupation: Unskilled, n (%) | 15 (30%) | 8 (16%) | — |
| Duration of illness/follow-up (months), mean ± SD | 19.26 ± 10.63 | 18.32 ± 10.32 | NS |

NS = not significant ( $p > 0.05$ by chi-square or independent t-test as appropriate).

Grouped bar charts showing the distribution of (A) sex, (B) residence, (C) marital status, and (D) educational level in MDD patients (blue) and healthy controls (green). No statistically significant between-group differences were found for any sociodemographic variable (all p > 0.05 by chi-square test).

### 3.2 Serum H₂S Levels

Serum H₂S concentrations were markedly and significantly lower in MDD patients compared with healthy controls (Table 2; Figure 4). The mean serum H₂S in the MDD group was 0.068 ± 0.044 µmol/L, compared with 0.524 ± 0.272 µmol/L in the control group, showing an approximately 7.7-fold reduction (p < 0.001).

**Figure 4:**
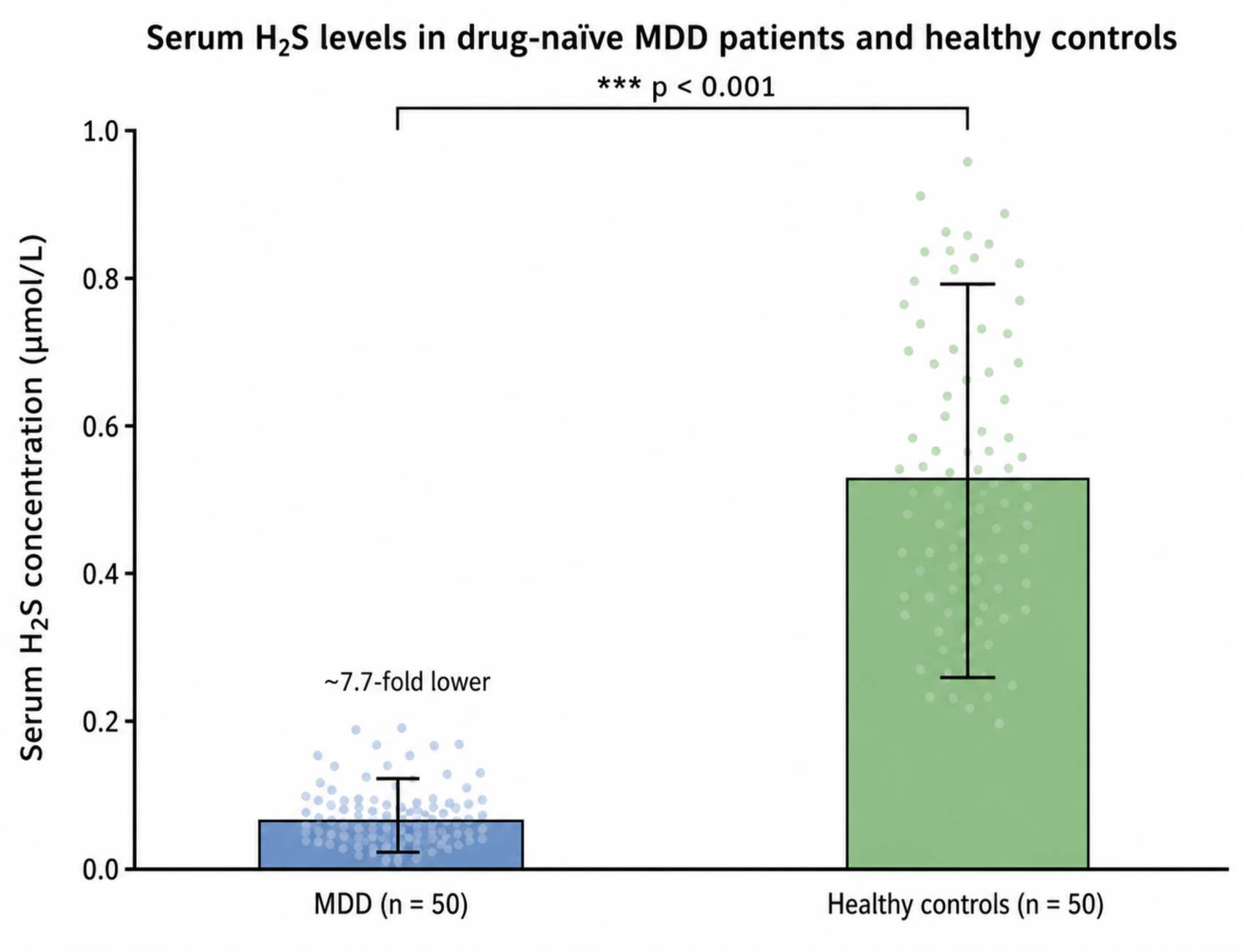
Serum H₂S levels in MDD patients and healthy controls.

**Table 2.**
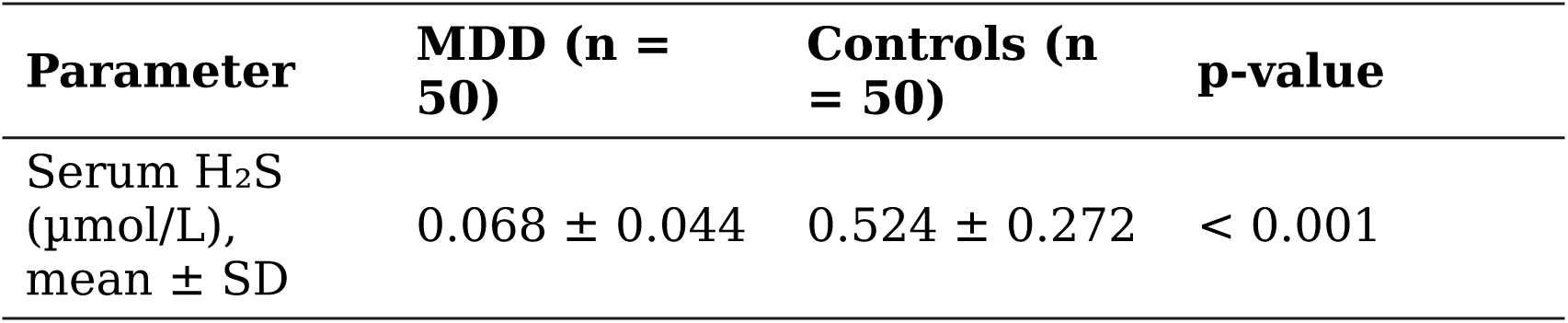
Comparison of serum H₂S between MDD patients and healthy controls.

| Parameter | MDD (n = 50) | Controls (n = 50) | p-value |
| --- | --- | --- | --- |
| Serum H<sub>2</sub>S (μmol/L), mean ± SD | 0.068 ± 0.044 | 0.524 ± 0.272 | < 0.001 |

#### Independent-samples t-test

The large SD in the control group reflected by 0.272 µmol/L relative to a mean of 0.524 µmol/L shows genuine biological variability in H₂S levels among healthy individuals, consistent with the known dependence of H₂S on dietary sulphur intake, gut microbiome composition, and CBS/CSE enzyme polymorphisms. The SD in the MDD group of 0.044 µmol/L relative to a mean of 0.068 µmol/L is proportionally smaller, suggesting a more uniform suppression of H₂S across the MDD group which is consistent with a disease-driven floor effect.

### 3.3 HAM-D-17 Scores

HAM-D-17 scores were significantly higher in MDD patients than in healthy controls (Table 3; Figure 5). The mean HAM-D-17 score in the MDD group was 28.94 ± 12.78, indicating predominantly severe to very severe depression (HAM-D ≥ 19), compared with a mean of 3.96 ± 2.31 in the control group (p < 0.001). The high mean HAM-D score in the MDD group is consistent with the tertiary care setting of this study, where patients with more severe illness are more likely to present.

**Figure 5:**
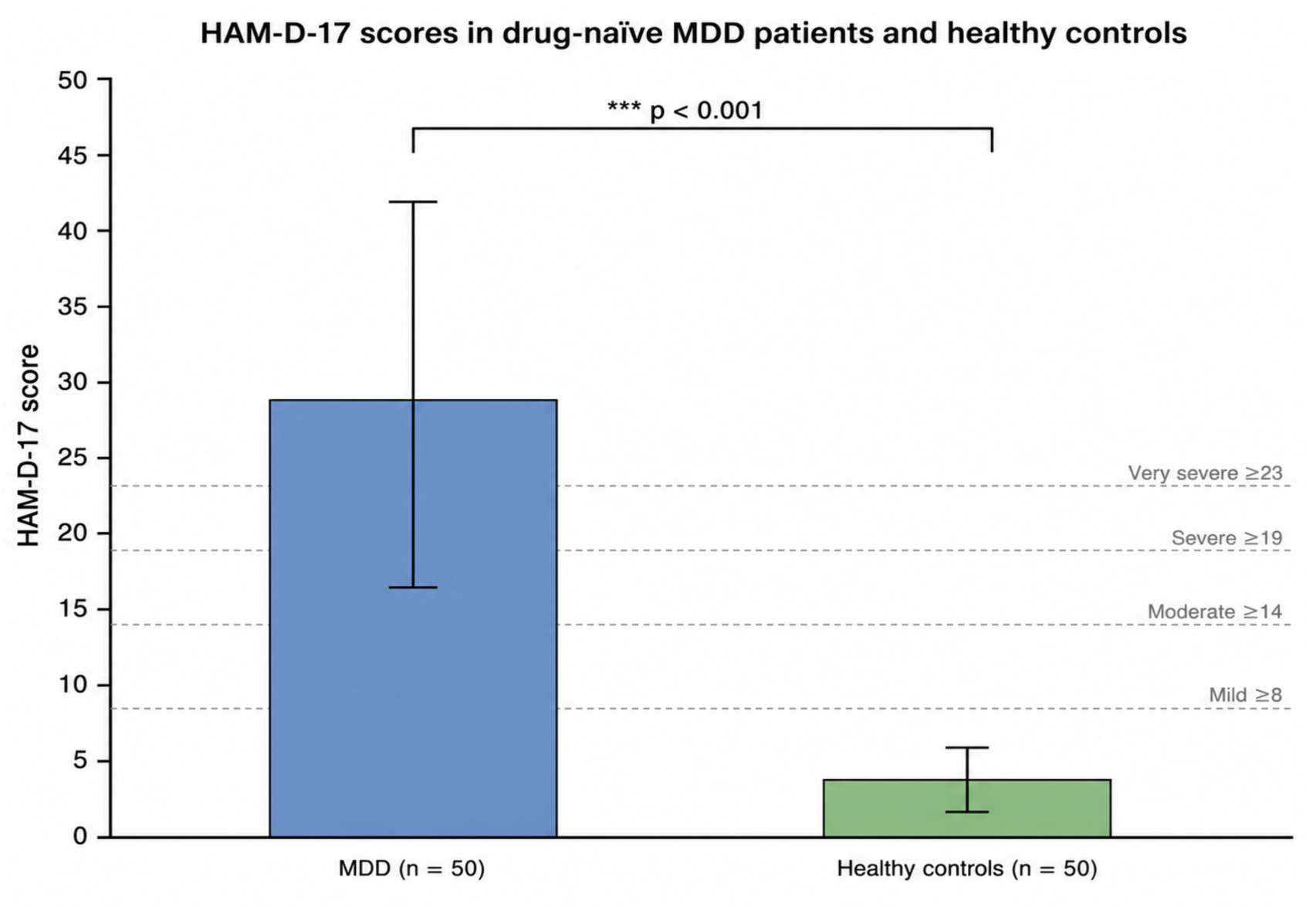
HAM-D-17 scores in MDD patients and healthy controls. Bar chart showing mean ± SD HAM-D-17 scores in MDD patients (n = 50; blue) and healthy controls (n = 50; green). Dashed horizontal lines indicate HAM-D severity thresholds (mild: 8–13; moderate: 14–18; severe: 19–22; very severe: ≥23). The mean HAM-D score in the MDD group (28.94 ± 12.78) falls in the very severe range. ***p < 0.001 (independent-samples t-test).

**Table 3.** Comparison of HAM-D-17 scores between MDD patients and healthy controls.

| Parameter | MDD (n = 50) | Controls (n = 50) | p-value |
| --- | --- | --- | --- |
| HAM-D-17 score, mean ± SD | $28.94 \pm 12.78$ | $3.96 \pm 2.31$ | $< 0.001$ |

### 3.4 Association Between Serum H₂S and HAM-D-17 Score

Linear regression analysis was performed to examine the relationship between serum H₂S (independent variable) and HAM-D-17 score (dependent variable). Results are summarised in Table 4 and illustrated in Figure 6.

**Figure 6:**
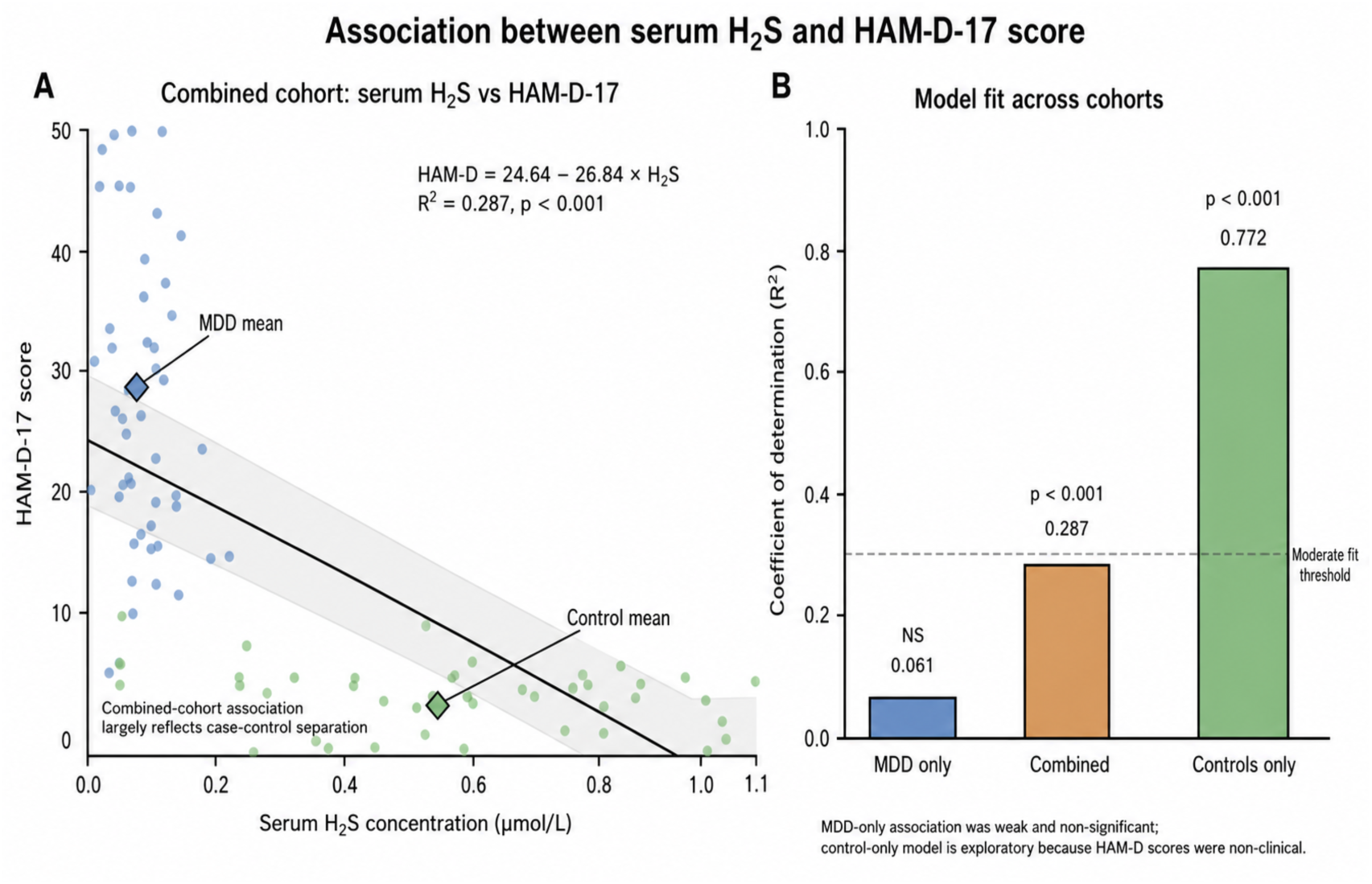
Association between serum H₂S and HAM-D-17 score. Panel A shows the combined-cohort regression between serum H₂S and HAM-D-17 score (R² = 0.287, p < 0.001), largely reflecting separation between MDD patients and healthy controls. Panel B compares regression model fit across cohorts, showing weak non-significant association within MDD patients (R² = 0.061) and exploratory control-only association (R² = 0.772).

**Table 4.** Linear regression: serum H₂S vs. HAM-D-17 score.

| Cohort | Regression equation | R<sup>2</sup> | p-value | Interpretation |
| --- | --- | --- | --- | --- |
| Combined (MDD + Control, n = 100) | HAM-D = 24.64 – 26.84 × H<sub>2</sub>S | 0.287 | < 0.001 | Significant inverse association, largely reflecting case-control separation. |
| MDD group only (n = 50) | — | 0.061 | NS | Weak, non-significant association. H<sub>2</sub>S did not reliably predict severity within MDD. |
| Control group only (n = 50) | — | 0.772 | < 0.001 | Exploratory association within non-clinical HAM-D range, not interpreted as depression severity. |

In the combined cohort (MDD patients + controls, n = 100), the regression equation was: **HAM-D score = 24.64 − 26.84 × [H₂S (µmol/L)].** In the combined cohort, serum H₂S showed a significant inverse association with HAM-D-17 score, indicating that participants with lower H₂S tended to have higher depressive symptom scores. However, this association largely reflects the separation between MDD patients and healthy controls rather than a continuous severity relationship within the MDD group. When MDD patients were analysed separately, the association between serum H₂S and HAM-D-17 score was weak and non-significant. Therefore, in this cohort, serum H₂S appears to distinguish MDD patients from healthy controls, but it should not be interpreted as a confirmed marker of depression severity among MDD patients.

### 3.5 Association Between Serum H₂S and Sociodemographic Variables

We wanted to see if background factors influenced H₂S levels, so we split the H₂S data at the median and ran chi-square tests. We found no statistically significant links between H₂S levels and any sociodemographic variables (all p > 0.05). This means that factors like age, sex, residence, language, religion, marital status, education, occupation, socioeconomic status, family type, and the duration of the illness had no measurable impact on H₂S levels in either the patient or the control group. Similarly, when we looked exclusively at the MDD group, a patient’s specific HAM-D severity category did not significantly align with their H₂S levels.

## 4. Discussion

### 4.1 Principal Findings

The principal finding of this study is a marked and statistically significant reduction in serum H₂S levels in drug-naïve MDD patients compared with healthy controls, with an approximately 7.7-fold difference in mean concentrations (0.068 ± 0.044 µmol/L vs. 0.524 ± 0.272 µmol/L; p < 0.001). Furthermore, across the combined cohort, higher serum H₂S was significantly associated with lower HAM-D-17 scores (R² = 0.287; p < 0.001). These findings provide direct clinical evidence from an Indian population supporting the H₂S deficiency hypothesis of depression and are consistent with a growing body of preclinical and emerging clinical evidence.

### 4.2 Consistency with Preclinical Evidence

Our findings are consistent with and extend a substantial body of preclinical evidence implicating H₂S deficiency in the neurobiology of depression. Luo et al. demonstrated in a CUMS mouse model that hippocampal H₂S and CBS expression are significantly reduced in depression, and that restoring H₂S via SAM treatment reverses synaptic ultrastructural deficits and ameliorates depression-like behaviours, accompanied by recovery of BDNF, SYN, and PSD-95 expression [7]. Our clinical finding of profoundly reduced serum H₂S in MDD patients mirrors these preclinical observations and suggests that the CBS/CSE–H₂S axis is similarly dysregulated in human depression. The neuroinflammatory dimension of H₂S deficiency in depression is further illuminated by the work of Hao et al., who showed that LPS-induced depression in mice is characterised by reduced plasma H₂S, increased hippocampal p-NF-κB, decreased hippocampal CA1 neuron counts, and reduced SYN and PSD-95 expression [8]. Administration of the novel H₂S donor BGS reversed all these changes, suppressing NF-κB activation and restoring synaptic plasticity markers [8]. This is mechanistically coherent with the broader literature showing that H₂S persulfidates the p65 subunit of NF-κB, shifting its transcriptional programme from pro-inflammatory to pro-survival [5]. The neuroinflammatory hypothesis of MDD supported by elevated pro-inflammatory cytokines in depressed patients may therefore be partly mediated by H₂S deficiency, which removes a key brake on NF-κB-driven neuroinflammation. The comprehensive systematic review by Pinna et al. (2025) synthesised 14 preclinical studies published between 2021 and 2025 and identified multiple converging molecular pathways through which H₂S deficiency contributes to depression [6]. These include the suppression of the PI3K/AKT/mTOR pathway impairing neurogenesis and anti-autophagy signaling, downregulation of Sirt1 and Sirt6, thus promoting neuroinflammation and ferroptosis, activation of the cGAS-STING pathway driving innate immune activation, induction of ER stress (GRP78/PERK/ATF4/GADD34 axis), suppression of CREB/BDNF signaling, and impairment of the hippocampal Warburg effect, therefore reducing the neuronal energy availability [6]. Our clinical finding of a 7.7-fold reduction in serum H₂S in MDD patients is consistent with the magnitude of H₂S depletion required to activate these pathways in preclinical models, and provides the human correlate of these mechanistic observations.

### 4.3 Consistency with Clinical Evidence

Our findings directly align with the only other dedicated clinical study on H₂S in depression. In 2021, Yang et al. measured plasma H₂S in 47 depressed patients and 51 healthy controls in China using a highly sensitive MBB fluorometric method [9]. They found significantly lower H₂S levels in the depressed group (0.59 ± 0.29 µmol/L) compared to the controls (1.02 ± 0.34 µmol/L; p < 0.001) [9]. They also noted a clear gradient based on illness severity.

Patients with mild depression averaged 0.84 µmol/L, moderate cases averaged 0.62 µmol/L, and severe cases dropped to 0.38 µmol/L (ANOVA p < 0.001) [9]. Ultimately, this revealed a strong negative correlation between H₂S levels and depression severity (Pearson r = −0.484; p = 0.001) [9]. Importantly, Yang et al. showed that taking antidepressants did not significantly alter H₂S levels [9]. This suggests the drop in H₂S is a core feature of the illness itself, not just a side effect of medication [9]. Our study independently backs this up, because we only evaluated drug-naïve patients, we proved that H₂S reduction occurs even in the complete absence of any prior psychotropic exposure. Closer to home, Dutta et al. (2024) reported significantly reduced serum H₂S in patients experiencing major depressive episodes linked to bipolar disorder [10]. They conducted their research at NBMCH in West Bengal, the exact same institution as our current study, using the same methylene blue spectrophotometric method [10].

Their work also highlighted a severity-dependent gradient and showed excellent diagnostic accuracy of AUC = 0.964 for diagnosing MDP or major depressive episodes in the context of bipolar disorder and AUC = 0.860 for identifying severe depression [10]. Our current study extends these findings to unipolar, drug-naïve MDD. We provide the first evidence that an H₂S deficiency is not limited to bipolar depression but is a distinct feature of unipolar MDD as well. When we look at these three independent clinical datasets together of Yang et al. from China, Dutta et al. focusing on bipolar depression in eastern India, and our own work on unipolar MDD in the same region, a clear picture emerges. The combined evidence strongly suggests that reduced circulating H₂S is a consistent biological marker for depressive illness, regardless of the patient population, the specific diagnostic subtype, or the measurement method used.

### 4.4 Interpretation of the Regression Findings

Our regression analysis conveys clearly that serum H₂S is a strong indicator of *whether* someone has depression, but it is not a precise dial that tracks *how severe* that depression is. When we looked at the entire group of participants together, we saw a significant trend that lower H₂S levels mapped clearly to higher HAM-D-17 scores. This aligns perfectly with the idea that H₂S plays a crucial role in protecting the brain. However, when we isolated just the MDD patients, that mathematical link vanished.

Because of this, we cannot safely say that measuring a patient’s H₂S level will accurately predict the day-to-day severity of their symptoms. We concluded that the link disappeared most likely due to a floor effect. The H₂S levels in our MDD patients were so universally depleted that they essentially hit biological rock bottom, leaving barely any variation to compare against their changing symptom scores. Depression is also deeply complex. A patient’s exact severity is driven by a web of factors far beyond just H₂S, including stress hormone pathways, inflammation, sleep disruption, and the sheer length of their illness. Low H₂S is a core piece of the depressive puzzle, but it is not the only thing controlling how bad the depression gets. We also need to treat the control-only data with caution. Healthy volunteers naturally have mood scores that cluster tightly in a narrow, non-clinical range.

With a smaller sample size, the strong correlation we saw in this healthy group might simply be statistical noise influenced by unmeasured lifestyle factors rather than a strict biological rule. Ultimately, this study establishes serum H₂S as a strong candidate marker for identifying the *presence* of MDD. To figure out if it can reliably track illness *severity*, the field will need larger, long-term studies that follow patients across a much wider spectrum of depression.

### 4.5 Absence of Sociodemographic Confounding

The absence of any significant association between serum H₂S and sociodemographic variables including sex, age, residence, education, occupation, and socioeconomic status in either group is an important finding. It suggests that the H₂S reduction observed in MDD is not confounded by these factors and may represent a relatively specific biological correlate of the depressive state rather than a reflection of social determinants of health. This is consistent with the finding of Yang et al. (2021), who similarly found no significant association between H₂S and demographic variables in their cohort [9].

### 4.6 Drug-Naïve Design as a Methodological Strength

The drug-naïve design of this study is a critical methodological strength. SAM, which is used as a nutraceutical antidepressant, acts in part by upregulating CBS and elevating H₂S [7]. Many conventional antidepressants may also indirectly modulate H₂S pathways through effects on oxidative stress, inflammation, and mitochondrial function. By restricting enrolment to drug-naïve patients, this study eliminates the confounding effect of psychotropic medication on H₂S levels and provides the clearest available evidence that H₂S reduction is an intrinsic feature of the depressive state rather than a pharmacological artefact. This is particularly important in the context of Yang et al.’s finding that antidepressant treatment did not significantly alter H₂S levels [9], which, while reassuring, does not exclude the possibility of treatment effects in other populations or with different antidepressant classes.

### 4.7 Therapeutic Implications

These findings open clear paths for treating depression. If future research confirms that a lack of H₂S is a core feature of MDD, we can start developing treatments that specifically target the CBS/ CSE–H₂S pathway. We actually already use some of these mechanisms today. For example, several countries use SAM as a nutritional antidepressant, and it works partly by boosting CBS activity to raise H₂S levels [7]. In laboratory settings, researchers are testing slow-release H₂S donors like GYY4137 and BGS, which have shown antidepressant-like effects in some animal models, though evidence remains inconsistent across studies [8].

Scientists are even designing highly specific compounds like AP39, a mitochondria-targeted H₂S donor, delivers H₂S directly to mitochondria, the exact part of the cell most vulnerable to oxidative stress during a depressive episode [5]. Because depression depletes both H₂S and GABA, dual-action treatments like BGS that restore both chemicals at the same time are especially promising [8]. We might also explore everyday ways to boost H₂S naturally. Patients could potentially take L-cysteine supplements, adjust their diets to include more sulfur, or use targeted probiotics to encourage their gut bacteria to produce sulfate [6]. That said, we need to be realistic about the timeline. As Pinna et al. recently pointed out, for depression in humans, meaning these targeted therapies are likely 10 to 15 years away from reaching the clinic [6].

### 4.8 Strengths, Limitations and Future Directions

Our study has several key strengths. First, we focused on a relatively unexplored gasotransmitter pathway in a strictly defined group of drug-naïve MDD patients. This allowed us to rule out the possibility that psychiatric medications caused the drop in H₂S. By including healthy controls and using a standardized biochemical assay, we were able to compare serum H₂S levels directly between the two groups. The massive difference we found strongly suggests that H₂S dysregulation is a meaningful biological marker for depression. However, we must view these findings with a few limitations in mind. Because our study is cross-sectional, we can only prove an association, not cause and effect.

We cannot say for certain if a drop in H₂S actually triggers depression, or if the depressive state itself causes H₂S levels to fall. To answer that, future researchers will need to track patients over time, measuring H₂S before and after treatment to see if levels bounce back as patients recover. Second, while our sample size was large enough to reveal a stark contrast between patients and controls, we conducted this study at a single hospital. To ensure these results apply to the broader population, we need larger, multi-center studies that span diverse demographic and clinical settings. Third, we used the widely accepted methylene blue method to measure serum H₂S. While reliable, this technique does not differentiate between free H₂S and other bound sulphide pools in the blood. Future research should use more precise tools like fluorometric probes, HPLC, or mass spectrometry to paint a more detailed picture of H₂S metabolism. Fourth, we only looked at serum H₂S and did not measure the actual activity of the enzymes that produce it (like CBS, CSE, or 3-MST), nor did we track related metabolites like sulphate or homocysteine. Adding these measurements in the future will help pinpoint exactly where the H₂S assembly line breaks down in MDD. Fifth, readers should interpret our absolute H₂S concentrations strictly within the context of our lab setup. Reported H₂S values vary wildly from lab to lab simply because of differences in calibration, sample handling, and detection equipment. Therefore, the real takeaway from our data isn’t the raw numbers themselves, but the massive relative drop in H₂S seen in depressed patients compared to the healthy controls. Finally, while looking at the combined cohort showed that lower H₂S links to worse depression, that mathematical trend vanished when we isolated just the MDD patients. Because of this, we can confidently say that H₂S is a great marker for *detecting* the presence of depression, but we need larger studies with a wider range of symptom severity to prove if it can accurately track *how bad* the depression is.

## 5. Conclusion

This cross-sectional study demonstrates that drug-naïve patients with Major Depressive Disorder have markedly reduced serum H₂S levels compared with healthy controls (∼7.7-fold reduction; p < 0.001), and that lower serum H₂S is significantly associated with greater depression severity as measured by HAM-D-17 (R² = 0.287; p < 0.001). These findings provide direct clinical evidence from an Indian population supporting the H₂S deficiency hypothesis of depression and are consistent with preclinical data demonstrating that H₂S depletion impairs synaptic plasticity, promotes neuroinflammation, and reduces BDNF signalling, as well as with the emerging clinical evidence from Yang et al. (2021) and Dutta et al. (2024). The absence of sociodemographic confounding and the drug-naïve design strengthen the inference that H₂S reduction is an intrinsic biological correlate of the depressive state. The CBS/CSE–H₂S axis warrants further investigation as a biomarker and therapeutic target in MDD. Longitudinal studies with larger samples, comprehensive H₂S speciation, and enzyme activity assays are needed to establish causality and to evaluate the antidepressant potential of H₂S-restoring strategies.

## Data Availability

All data produced in the present study are available upon reasonable request to the authors

## References

1. World Health Organization. Depressive disorder (depression). WHO Fact Sheet. 2023. Available at: https://www.who.int/news-room/fact-sheets/detail/depression

2. Rush AJ, Trivedi MH, Wisniewski SR, et al. Acute and longer-term outcomes in depressed outpatients requiring one or several treatment steps: a STAR\**D report*. Am J Psychiatry. 2006;163(11):1905–1917. doi:10.1176/ajp.2006.163.11.1905

3. Malhi GS, Mann JJ. Depression. Lancet. 2018;392(10161):2299–2312. doi:10.1016/S0140-6736(18)31948-2

4. Wang R. Two’s company, three’s a crowd: can H₂S be the third endogenous gaseous transmitter? FASEB J. 2002;16(13):1792–1798. doi:10.1096/fj.02-0211hyp

5. Morris JL, Lee JJ, Morris RE, Miljkovic JLj. Black gas, bright future: H₂S based therapeutics for neurodegenerative disorders. Neurotherapeutics. 2025;22:e00755. doi:10.1016/j.neurot.2025.e00755

6. Pinna A, Kistowska J, Pałasz A. An enigma of brain gasotransmitters: hydrogen sulfide and depression. NeuroMolecular Medicine. 2025;27:58. doi:10.1007/s12017-025-08880-y

7. Luo Y, Yue W, Quan X, Wang Y, Zhao B, Lu Z. S-Adenosyl methionine improves depression-like behaviours and synaptic markers by elevating the expression of endogenous hydrogen sulfide in the hippocampus. Neuropsychiatry. 2018;8(2):495–504.

8. Hao X, Wang C, Liu S, Yu P, Liu Y, Ma Y. Design and synthesis of a multifunctional hydrogen sulfide donor and its application in depressive-like behavior in mice induced by lipopolysaccharide. ACS Chemical Neuroscience. 2025;16:4267–4278. doi:10.1021/acschemneuro.5c00579

9. Yang J, Liang J, Hu N, et al. A new clinical study and meta-analysis reveals decreased plasma hydrogen sulfide levels in patients with depression. Frontiers in Psychiatry. 2021;12:765664. doi:10.3389/fpsyt.2021.765664

10. Dutta S, Bhattacharya S, Mondal S, et al. Serum hydrogen sulfide as a biomarker of depression severity in patients with major depressive episodes: a case-control study. Cureus. 2024;16(3):e56857. doi:10.7759/cureus.56857

11. American Psychiatric Association. Diagnostic and Statistical Manual of Mental Disorders, 5th ed. (DSM-5). Washington, DC: APA; 2013.

12. Hamilton M. A rating scale for depression. J Neurol Neurosurg Psychiatry. 1960;23:56–62. doi:10.1136/jnnp.23.1.56

13. Kuppuswamy B. Manual of Socioeconomic Status (Urban). Delhi: Manasayan; 1981.

